# Early Diagnosis of Cerebral Palsy in Preterm Infants with MRI, General Movements and Neurological Exam

**DOI:** 10.1101/2024.12.10.24318810

**Authors:** Shipra Jain, Karen Harpster, Stephanie Merhar, Beth Kline-Fath, Mekibib Altaye, Venkata Sita Priyanka Illapani, Colleen Peyton, Nehal A. Parikh, Cincinnati Infant Neurodevelopment Early Prediction Study (CINEPS) Investigators

**Author notes:** Corresponding Author: Nehal A. Parikh, DO, MS, Professor of Pediatrics, Cincinnati Children’s Hospital Medical Center, 3333 Burnett Avenue, Cincinnati, Ohio, 45229, United States.

## Abstract

**Background:** The increasing clinical use of combining structural MRI (sMRI) with General Movements Assessment (GMA) or Hammersmith Infant Neurological Exam (HINE) before five months corrected age (CA) for early diagnosis of cerebral palsy (CP) lacks sufficient prognostic data for children with CP, especially those with Gross Motor Function Classification System (GMFCS) I.

**Objective:** Evaluate the predictive value of sMRI, GMA, and HINE individually and in combination for early CP diagnosis and assess accuracy across varying GMFCS levels in a regional cohort of preterm infants.

**Methods:** We performed sMRI between 39-44 weeks postmenstrual age and GMA and HINE between 12-18 weeks CA in 395 preterm infants born at ≤32 weeks’ gestation across five NICUs in Greater Cincinnati. Brain abnormalities on sMRI included white matter injuries, cortical and deep gray matter lesions, or extensive cerebellar hemorrhage. Absent fidgety movements constituted abnormal GMA; abnormal HINEs were scores <56. The primary outcome was CP diagnosis at 22-26 months CA, classified by the GMFCS. We calculated sensitivity, specificity, positive predictive value, negative predictive value, and likelihood ratios for individual tests and combinations.

**Results:** Of 338 (86%) infants with complete follow-up, 48 (14.2%) showed sMRI abnormalities, 15 (4.6%) had abnormal GMA, and 69 (20.9%) had abnormal HINE. Thirty-nine children (11.5%) developed CP at age 2, of which 28 had GMFCS level I and 11 had GMFCS ≥II. The combination of sMRI and GMA achieved 100% specificity but only 22% sensitivity while the combination of abnormal sMRI and HINE demonstrated sensitivity of 32% and specificity of 98% for prediction of CP. Individual or combined tests showed far higher sensitivity (78-100%) for predicting CP in children with GMFCS levels II-V.

**Conclusions:** The combination of sMRI with GMA or HINE demonstrated high specificity but low sensitivity for early CP diagnosis in a regional cohort of preterm infants. This approach appears effective for early detection of CP levels II-V but not for level I cases, the most prevalent type, underscoring the need for continued developmental follow-up for all very preterm infants and need for more sensitive diagnostic tools for early detection of CP.

**Key Points:** *Questions:* What is the individual and combined prognostic accuracy of sMRI, GMA, and HINE for early diagnosis of CP in preterm infants?

*Findings:* In our prospective, regional study of preterm infants born at ≤32 weeks’ gestation, we found that combining brain abnormalities on sMRI with abnormal GMA achieved 100% specificity but 22% sensitivity for diagnosing CP. Individual or combined tests showed far higher sensitivity (78-100%) for predicting CP in children with GMFCS levels II-V. Both individual and combined tests were poor predictors of GMFCS level I CP, the most common type.

*Meaning:* While sMRI combined with GMA or HINE is effective for diagnosing CP with GMFCS levels II-V, this approach falls short for children with GMFCS level I.

## Introduction

Cerebral palsy (CP) affects 0.2-0.3% of live births in the United States,^1,2^ with risk increasing to 6-18% in preterm infants, particularly those born at ≤32 weeks gestational age (GA).^1,3,4^ CP is a movement and posture disorder caused by brain injury or abnormalities, with outcomes ranging from independent ambulation to complete dependence for all mobility.^5^ While a definitive cure for CP remains elusive, early intervention can curtail the progression of motor deficits, prevent secondary impairments, and enhance independence.^6–8^ The brain exhibits its highest levels of neuroplasticity during early life,^9^ underscoring the importance of early CP diagnosis and subsequent early intervention for improved prognosis.

Prognostic/diagnostic tools for detecting CP in infants below 5 months corrected age (CA) include structural magnetic resonance imaging (sMRI) at term-equivalent age (TEA), Prechtl’s General Movements Assessment (GMA) during the fidgety period, and Hammersmith Infant Neurological Examination (HINE) at 3-4 months CA.^10–12^ These tests individually lack sufficient predictive value for accurate CP diagnosis, and their combined effectiveness remains limited.^13–15^ A recent consensus recommended combining TEA-sMRI with GMA or HINE between 3-5 months CA for early diagnosis in high-risk newborns.^16^ While a retrospective case-control study reported 98% sensitivity and 99% specificity when combining neuroimaging (ultrasound or sMRI), GMA, and HINE^14^, only one cohort study (N=130) in very preterm infants assessed the prognostic value of combining sMRI with GMA, yielding positive predictive value (PPV) of 35% (sensitivity 70%; specificity 82%), falling short of previously reported findings and recommendations.^15^

The above guideline stated prognostic values from prior systematic reviews that reported sensitivity between 86%-89% for sMRI and 98% for GMA in predicting CP.^16^ However, these results should be applied cautiously to preterm infants. Many studies in the review^12,14,16–24^ focused on high-risk infants with perinatal asphyxia and stroke, where CP prevalence is much higher (6.3%–52.4%), potentially introducing bias and increasing the tests’ prognostic accuracy. A large sMRI study (N=480), not included in the review, demonstrated a much lower sMRI sensitivity of 48% and a PPV of 17% for CP prediction.^25^ Likewise, external validation studies by advanced GMA-trained practitioners revealed lower GMA sensitivity (50%–56%).^15,26^ Furthermore, predictive accuracy across different Gross Motor Functional Classification Scale (GMFCS) levels of CP remains underexplored.

To address these gaps, we prospectively evaluated the prognostic value of combining TEA- sMRI with GMA or HINE at 3-4 months CA for early CP diagnosis and assessed the predictive accuracy of these tools individually and in combination for varying CP GMFCS levels in preterm infants.

## Methodology

### Study Design and Participants

Our multisite, prospective Cincinnati Infant Neurodevelopment Early Prediction Study (CINEPS) included 395 infants born at ≤32 weeks GA across five level III/IV neonatal intensive care units in Greater Cincinnati between 09/2016 to 11/2019, excluding infants with known congenital or chromosomal anomalies affecting the central nervous system or those still hospitalized at 44 weeks postmenstrual age (PMA), except those at Cincinnati Children’s Hospital Medical Center, the imaging site. The Institutional Review Board approved the study, and written parental/guardian consents were obtained.

### Structural MRI (sMRI) acquisition and assessment

We acquired high-resolution 3 Tesla T2- weighted, 3D T1-weighted, and susceptibility weighted MRI sequences at 39-44 weeks PMA to detect brain abnormalities (Supplement 1).^27^ One neuroradiologist (B.M.K.F.), blinded to clinical and outcome data, evaluated scans with high intra-rater reliability.^28^ sMRI was considered abnormal if it showed white matter injuries (e.g., cystic periventricular leukomalacia), encephalomalacia, periventricular hemorrhagic infarctions, lesions in the cortex or basal ganglia/thalamus, or extensive cerebellar hemorrhage—predictive abnormalities for CP, as outlined by Novak et al.^16^

### Neurodevelopmental outcome assessment

General Movement Assessment (GMA): GMA categorizes spontaneous movements in alert, quiet, infants. Infants between 12-18 weeks CA were video recorded and scored by an advanced Prechtl-certified examiner (K.H.), masked to clinical history and sMRI findings. ^29,30^ The same examiner assessed the intra-rater reliability by re-evaluating 30 randomly selected videos 6 months later. Infants with normal fidgety movements exhibited variable, complex, and multiplanar movements, while those with absent or sporadic (≤3 seconds) fidgety movements were classified as having abnormal GMA, predictive of motor dysfunction, particularly CP.^30^ No cohort infants exhibited abnormal fidgety movements (exaggerated amplitude or speed).^31,32^ A General Movements Trust trainer (C.P.) provided secondary assessment for equivocal videos.

Hammersmith Infant Neurological Examination (HINE): HINE is a standardized neurologic examination for infants aged 2-24 months CA evaluating infants’ posture, tone, cranial nerves, reflexes, and spontaneous and voluntary movements, scoring infants on a 0–78 scale. ^24^ Scores <56 were considered abnormal.^33^ One examiner (K.H.), masked to clinical history and MRI findings, performed HINEs on the same day as GMAs. The same examiner assessed the intra-rater reliability by retesting 15 randomly selected infants 2-4 weeks after their initial HINE.

Neurological examination to determine CP diagnosis: Certified examiners^34^ performed the Amiel-Tison standardized neurological exam^35^ at 22-26 months CA to assess motor function and diagnose CP using Neonatal Research Network criteria: (1) abnormalities in tone, deep tendon reflexes, coordination, and/or movement, except isolated hypotonia or toe walking without tight ankles; (2) delays in motor milestones with motor dysfunction and (3) possible aberrations in reflexes and postural reactions. Because this definition permits a diagnosis of level I CP for a child with isolated toe walking with tight ankles, we required at least one additional delay in milestones or tone abnormalities for CP diagnosis. We used the GMFCS to rate CP gross motor functional ability on a scale of I to V.^36,37^ Our primary outcome was CP diagnosis at 22-26 months CA.

### Statistical analysis

We compared baseline differences between infants with and without CP and those who returned for follow-up at 2 years CA vs. those who did not, using Student’s t-test, Mann Whitney U, Chi-Squared, or Fisher’s exact test as appropriate. Intra-rater reliability was assessed using kappa statistics for dichotomous variables and intraclass correlation coefficient (ICC) for continuous variables. Fisher’s exact test examined the sensitivity, specificity, PPV, and negative predictive value (NPV) of the three tests individually and in combination for CP diagnosis at 22-26 months CA. To account for disease prevalence between centers, we calculated likelihood ratios (LR+ and LR-) for positive and negative tests. Logistic regression determined the odds ratio for CP for individual and combined tests. Predicted post-test probabilities were derived using the *margins* command for different GMFCS levels in children with CP for each test and their combinations. Analysis was performed using Stata 17.0, with two-sided P values <0.05 considered significant.

## Results

Of 395 enrolled infants, two withdrew, and one died before age 2. At 12-18 weeks CA, 381 (97%) returned for GMA and HINE testing, and 338 (86%) underwent neurological examinations for CP diagnosis at 22-26 months CA. The mean (SD) GA was 29.3 (2.5) weeks, and the mean birth weight (SD) was 1294 (449) grams. Table 1 represents the demographic characteristics of infants with and without CP. The demographic characteristics of infants who returned for the 2-year evaluation versus those who did not, differed in their GA (29 weeks, 3 days with follow-up versus 30 weeks, 0 days without follow-up, p=0.02) and race/ethnicity, p=0.003.

**Table 1:**
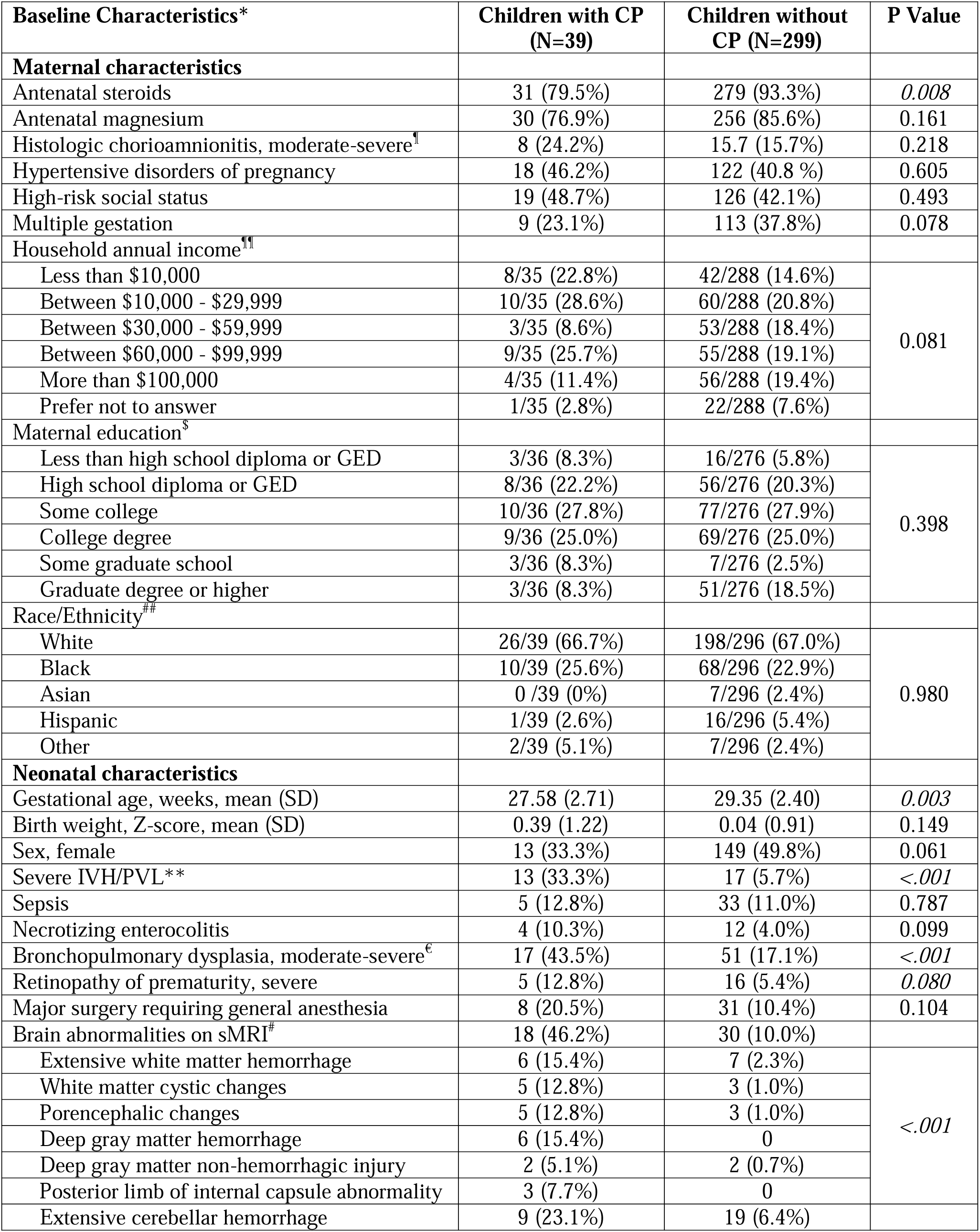
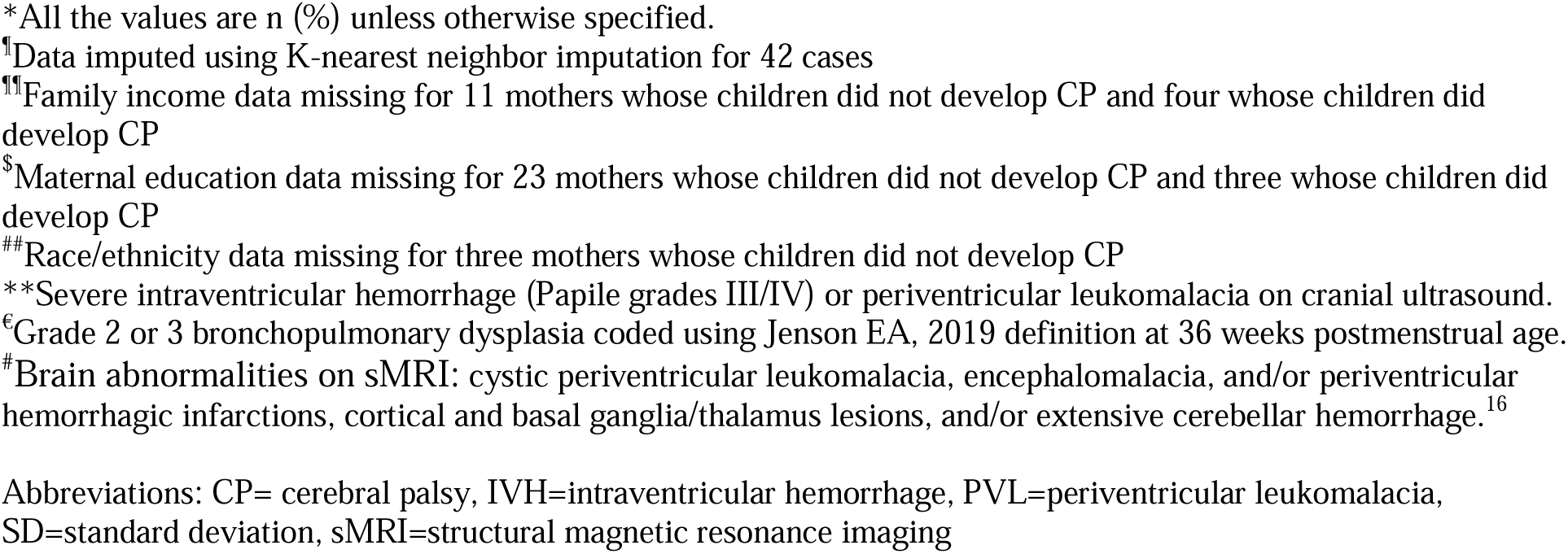
Baseline maternal and infant characteristics of children with and without cerebral palsy (CP)

| Baseline Characteristics* | Children with CP<br>(N=39) | Children without CP (N=299) | P Value |
| --- | --- | --- | --- |
| **Maternal characteristics** |  |  |  |
| Antenatal steroids | 31 (79.5%) | 279 (93.3%) | 0.008 |
| Antenatal magnesium | 30 (76.9%) | 256 (85.6%) | 0.161 |
| Histologic chorioamnionitis, moderate-severe<sup>†</sup> | 8 (24.2%) | 15.7 (15.7%) | 0.218 |
| Hypertensive disorders of pregnancy | 18 (46.2%) | 122 (40.8 %) | 0.605 |
| High-risk social status | 19 (48.7%) | 126 (42.1%) | 0.493 |
| Multiple gestation | 9 (23.1%) | 113 (37.8%) | 0.078 |
| Household annual income<sup>††</sup> |  |  |  |
| Less than \$10,000 | 8/35 (22.8%) | 42/288 (14.6%) | 0.081 |
| Between \$10,000 - \$29,999 | 10/35 (28.6%) | 60/288 (20.8%) | |
| Between \$30,000 - \$59,999 | 3/35 (8.6%) | 53/288 (18.4%) | |
| Between \$60,000 - \$99,999 | 9/35 (25.7%) | 55/288 (19.1%) | |
| More than \$100,000 | 4/35 (11.4%) | 56/288 (19.4%) | |
| Prefer not to answer | 1/35 (2.8%) | 22/288 (7.6%) |  |
| Maternal education<sup>§</sup> |  |  |  |
| Less than high school diploma or GED | 3/36 (8.3%) | 16/276 (5.8%) | 0.398 |
| High school diploma or GED | 8/36 (22.2%) | 56/276 (20.3%) |  |
| Some college | 10/36 (27.8%) | 77/276 (27.9%) |  |
| College degree | 9/36 (25.0%) | 69/276 (25.0%) |  |
| Some graduate school | 3/36 (8.3%) | 7/276 (2.5%) |  |
| Graduate degree or higher | 3/36 (8.3%) | 51/276 (18.5%) |  |
| Race/Ethnicity<sup>##</sup> |  |  |  |
| White | 26/39 (66.7%) | 198/296 (67.0%) | 0.980 |
| Black | 10/39 (25.6%) | 68/296 (22.9%) |  |
| Asian | 0 /39 (0%) | 7/296 (2.4%) |  |
| Hispanic | 1/39 (2.6%) | 16/296 (5.4%) |  |
| Other | 2/39 (5.1%) | 7/296 (2.4%) |  |
| **Neonatal characteristics** |  |  |  |
| Gestational age, weeks, mean (SD) | 27.58 (2.71) | 29.35 (2.40) | 0.003 |
| Birth weight, Z-score, mean (SD) | 0.39 (1.22) | 0.04 (0.91) | 0.149 |
| Sex, female | 13 (33.3%) | 149 (49.8%) | 0.061 |
| Severe IVH/PVL** | 13 (33.3%) | 17 (5.7%) | <.001 |
| Sepsis | 5 (12.8%) | 33 (11.0%) | 0.787 |
| Necrotizing enterocolitis | 4 (10.3%) | 12 (4.0%) | 0.099 |
| Bronchopulmonary dysplasia, moderate-severe<sup>€</sup> | 17 (43.5%) | 51 (17.1%) | <.001 |
| Retinopathy of prematurity, severe | 5 (12.8%) | 16 (5.4%) | 0.080 |
| Major surgery requiring general anesthesia | 8 (20.5%) | 31 (10.4%) | 0.104 |
| Brain abnormalities on sMRI<sup>#</sup> | 18 (46.2%) | 30 (10.0%) |  |
| Extensive white matter hemorrhage | 6 (15.4%) | 7 (2.3%) | <.001 |
| White matter cystic changes | 5 (12.8%) | 3 (1.0%) |  |
| Porencephalic changes | 5 (12.8%) | 3 (1.0%) |  |
| Deep gray matter hemorrhage | 6 (15.4%) | 0 |  |
| Deep gray matter non-hemorrhagic injury | 2 (5.1%) | 2 (0.7%) |  |
| Posterior limb of internal capsule abnormality | 3 (7.7%) | 0 |  |
| Extensive cerebellar hemorrhage | 9 (23.1%) | 19 (6.4%) |  |

\*All the values are n (%) unless otherwise specified.
<sup>¶</sup>Data imputed using K-nearest neighbor imputation for 42 cases
<sup>¶¶</sup>Family income data missing for 11 mothers whose children did not develop CP and four whose children did develop CP
<sup>\$</sup>Maternal education data missing for 23 mothers whose children did not develop CP and three whose children did develop CP
<sup>##</sup>Race/ethnicity data missing for three mothers whose children did not develop CP
<sup>\*\*</sup>Severe intraventricular hemorrhage (Papile grades III/IV) or periventricular leukomalacia on cranial ultrasound.
<sup>€</sup>Grade 2 or 3 bronchopulmonary dysplasia coded using Jenson EA, 2019 definition at 36 weeks postmenstrual age.
<sup>#</sup>Brain abnormalities on sMRI: cystic periventricular leukomalacia, encephalomalacia, and/or periventricular hemorrhagic infarctions, cortical and basal ganglia/thalamus lesions, and/or extensive cerebellar hemorrhage.<sup>16</sup>
Abbreviations: CP= cerebral palsy, IVH=intraventricular hemorrhage, PVL=periventricular leukomalacia, SD=standard deviation, sMRI=structural magnetic resonance imaging

We performed sMRI at a mean PMA of 42.3 (SD 1.3) weeks; 48 of 338 infants (14.2%) showed brain injury/abnormalities. GMA was completed for 329 infants between 12-18 weeks CA, with 97% conducted within the optimal 12-16 weeks CA. Fifteen (4.6%) infants had absent fidgety movements. HINE scores, available for 331 infants, ranged from 30.5 to 73 (median 60, IQR 7.0), with abnormal scores (<56) in 69 (20.9%) infants. Intra-rater reliability for GMA was 96.7% (kappa 0.89). Intra-rater ICC for HINE was 0.79.

At 22-26 months CA, 39 (11.5%) children were diagnosed with CP: 28 (8.3%) with GMFCS level I, 6 (1.8%) with GMFCS levels II-III, and 5 (1.5%) with GMFCS levels IV-V.^24^Among these 39 children diagnosed with CP, sMRI was abnormal in 18 (46.2%), GMA was abnormal in 10 (27.0%; two with missing GMA), and HINE was abnormal in 22 (57.9%; one with missing HINE) (Table 2; Table 3). Among 299 children without CP, 30 had abnormalities on their sMRI, 5 on GMA, and 47 on their HINE (Table 2). Only 8 infants had abnormal sMRI and GMA, and 17 infants had abnormal sMRI and HINE (Table 2; Table 3).

**Table 2.** Frequency of abnormalities on individual and combination prognostic tests in children with and without cerebral palsy (CP)

| Test(s)* | Children with CP<br>(N=39) | Children without CP<br>(N=299) | P Value |
| --- | --- | --- | --- |
| Abnormal sMRI | 18/39 (46.2%) | 30/299 (10.0%) | <0.001 |
| Abnormal GMA | 10/37 (27.0%) | 5/292 (1.7%) | <0.001 |
| Abnormal HINE | 22/38 (57.9%) | 47/293 (16.0%) | <0.001 |
| HINE Score, median<br>(range) | 55 (30.5 - 71) | 60 (38.5 – 73) | <0.001 |
| Abnormal sMRI and<br>GMA | 8/37 (21.6%) | 0/292 (0%) | <0.001 |
| Abnormal sMRI and<br>HINE | 12/38 (31.6%) | 5/293 (1.7%) | <0.001 |
| Abnormal sMRI and<br>GMA or HINE | 12/38 (31.6%) | 5/293 (1.7%) | <0.001 |
| Bayley Motor score,<br>median (range) | 73 (46 – 112) | 94 (55-130) | <0.001 |

\*All values are n (%) unless otherwise noted
Abbreviations: CP= cerebral palsy, sMRI=structural magnetic resonance imaging, GMA=general movement assessment, HINE= Hammersmith Infant Neurological Examination

**Table 3.** Abnormality on structural MRI (sMRI), general movements assessment (GMA), and/or Hammersmith Infant Neurological Exam (HINE) for all 39 infants diagnosed with cerebral palsy at age 2 years corrected age and associated Gross Motor Functional Classification Scale (GMFCS) in the cohort.

| sMRI Abnormality | GMA Abnormal | HINE Abnormal | HINE Score | CP at 2 Years | GMFCS Level |
| --- | --- | --- | --- | --- | --- |
| No | No | No | 71 | Yes | I |
| No | No | Yes | 55 | Yes | I |
| Yes | Yes | Yes | 42 | Yes | I |
| Yes | No | No | 64 | Yes | IV |
| No | No | No | 63.5 | Yes | I |
| Yes | No | No | 64.5 | Yes | I |
| No | Yes | Yes | 52 | Yes | III |
| No | No | No | 57 | Yes | I |
| Yes | Yes | Yes | 51 | Yes | III |
| No | No | No | 61 | Yes | I |
| No | No | Yes | 30.5 | Yes | I |
| No | No | No | 56 | Yes | I |
| Yes | No | No | 64 | Yes | I |
| No | Yes | Yes | 55 | Yes | I |
| Yes | No | No | 63 | Yes | I |
| No | No | Yes | 41 | Yes | I |
| Yes | No | Yes | 51.5 | Yes | I |
| Yes | Yes | Yes | 30.5 | Yes | V |
| No | No | No | 61 | Yes | I |
| Yes | No | Yes | 36 | Yes | III |
| No | No | Yes | 55 | Yes | I |
| Yes | Yes | Yes | 52 | Yes | III |
| Yes | Yes | Yes | 37 | Yes | IV |
| Yes | No | Yes | 49.5 | Yes | I |
| Yes | Yes | Yes | 32 | Yes | V |
| Yes | Yes | Yes | 42.5 | Yes | V |
| No | No | No | 63 | Yes | I |
| No | No | Yes | 54.5 | Yes | I |
| No | No | No | 62.5 | Yes | I |
| Yes | Missing | Missing | Missing | Yes | II |
| No | No | No | 58 | Yes | I |
| No | No | No | 59.5 | Yes | I |
| Yes | No | No | 63 | Yes | I |
| No | No | Yes | 54 | Yes | I |
| No | No | No | 65 | Yes | I |
| No | No | Yes | 50 | Yes | I |
| Yes | Yes | Yes | 53.5 | Yes | I |
| No | No | Yes | 55 | Yes | I |
| Yes | Missing | Yes | 45 | Yes | II |
| 18/39 (46.2%) | 10/37 (27.0%) | 22/38 (57.9%) | 55 (49.5-62.5)* | 39/39 (100%) | N/A |

\*Median (IQR)

Table 3 represents the association between CP diagnosis and abnormal sMRI, GMA, and HINE. Infants with abnormal GMA exhibited the highest odds of developing CP at age 2 as compared to those with sMRI brain abnormalities or abnormal HINE. Combining abnormal sMRI with abnormal GMA resulted in 100% CP specificity, while combining sMRI with abnormal HINE increased the odds of developing CP from 7.7 to 26.6 (Table 4).

**Table 4.** Odds of abnormal structural MRI (sMRI), general movement assessment (GMA), and Hammersmith Infant Neurological Examination (HINE) and prediction of cerebral palsy (CP) at 2 years corrected age in preterm infants.

| Outcomes | Odds Ratio | P value |
| --- | --- | --- |
| **Any CP** |  |  |
| sMRI | 7.7 (3.7, 16.0) | <0.001 |
| GMA | 21.3 (6.8, 66.7) | <0.001 |
| HINE | 7.2 (3.5, 14.7) | <0.001 |
| sMRI plus GMA* | N/A | N/A |
| sMRI plus HINE<sup>†</sup> | 26.6 (8.7, 81.3) | <0.001 |
| **CP GMFCS levels II-V<sup>#</sup>** |  |  |
| sMRI | 76.1 (9.5, 610.8) | <0.001 |
| GMA | 357.7 (39.3, 3259.8) | <0.001 |
| HINE** | N/A | N/A |
| sMRI plus GMA | 1116.5 (90.3, 13801.2) | <0.001 |
| sMRI plus HINE<sup>†</sup> | 352.1 (39.7, 3121.0) | <0.001 |
| **CP GMFCS level I<sup>#</sup>** |  |  |
| sMRI | 2.7 (1.1, 6.5) | 0.028 |
| GMA | 1.7 (0.4, 8.0) | 0.498 |
| HINE | 3.2 (1.5, 7.2) | 0.004 |
| sMRI plus GMA | 1.6 (0.18, 13.1) | 0.685 |
| sMRI plus HINE<sup>†</sup> | 2.5 (0.7, 9.2) | 0.175 |

\*Predicts success perfectly due to 100% specificity
\*\*Predicts success perfectly due to 100% sensitivity
<sup>†</sup>Results for sMRI and GMA OR sMRI and HINE were identical to this group
<sup>#</sup> GMFCS Level I is defined as an infant's ability to move in and out of sitting and floor sit with both hands free to manipulate objects. Infants crawl on hands and knees, pull to stand and take steps holding on to furniture. Infants walk 10 steps independently, with hands free, but with some gait abnormalities – includes toe walking, asymmetric walking, wide based gait with coordination or ataxic gait. Level II to V are characterized by increasing levels of limitations to walking and voluntary control of movement in other muscles.<sup>36,37</sup>
Abbreviations: CP= cerebral palsy, sMRI=structural magnetic resonance imaging, GMA=general movement assessment, HINE= Hammersmith Infant Neurological Examination, GMFCS= Gross Motor Functional Classification Scale

Infants with level II-V CP at age 2 had significantly higher odds of abnormal sMRI, GMA, and HINE, individually and in combination (Table 4). However, only brain abnormalities on sMRI and abnormal HINE individually significantly predicted level I CP with an odds ratio of 2.7 (95% CI: 1.1, 6.5) and 3.2 (1.5, 7.2), respectively. Neither abnormal GMA nor the combination approaches were predictive of level I CP.

Table 5 illustrates the individual and combined prognostic accuracy of sMRI, GMA, and HINE in predicting CP at age 2. Tests performed well individually or in combination in identifying/ruling in CP but performed poorly in ruling out CP. Combining sMRI with GMA or HINE enhanced specificity but greatly reduced sensitivity (from 46% for sMRI alone to 22% when combined with GMA). For levels II-V CP, all tests individually and in combination were effective in ruling in the diagnosis, but sensitivity, while higher than for level I CP, remained suboptimal. The sMRI and GMA combination performed best with a PPV of 88% and NPV of 99% for predicting levels II-V CP but performed poorly at predicting level I CP (Table 5).

**Table 5:** Prognostics accuracy of structural MRI (sMRI), general movement assessment (GMA), and Hammersmith Infant Neurological Examination (HINE) for prediction of cerebral palsy (CP) at 2 years corrected age in preterm infants.

| **Prognostic Models** | **Sensitivity** | **Specificity** | **PPV** | **NPV** | **LR+** | **LR-** | **AUC** |
| --- | --- | --- | --- | --- | --- | --- | --- |
| **Any CP** |  |  |  |  |  |  |  |
| sMRI | 0.46 | 0.90 | 0.38 | 0.93 | 4.6 | 0.60 | 0.681 |
| GMA | 0.27 | 0.98 | 0.67 | 0.91 | 15.8 | 0.74 | 0.627 |
| HINE | 0.58 | 0.84 | 0.32 | 0.94 | 3.6 | 0.50 | 0.709 |
| sMRI plus GMA | 0.22 | 1.00 | 1.00 | 0.89 | ∞ | 0.78 | N/A* |
| sMRI plus HINE | 0.32 | 0.98 | 0.72 | 0.92 | 18.5 | 0.70 | 0.649 |
| **CP GMFCS levels II-V<sup>#</sup>** |  |  |  |  |  |  |  |
| sMRI | 0.91 | 0.88 | 0.21 | 0.997 | 7.8 | 0.10 | 0.896 |
| GMA | 0.89 | 0.98 | 0.53 | 0.997 | 40.6 | 0.11 | 0.934 |
| HINE | 1.00 | 0.82 | 0.15 | 1.00 | 5.4 | 0 | N/A** |
| sMRI plus GMA | 0.78 | 0.997 | 0.88 | 0.99 | 248.9 | 0.22 | 0.887 |
| sMRI plus HINE | 0.90 | 0.98 | 0.53 | 0.997 | 36.1 | 0.10 | 0.938 |
| **CP GMFCS level I<sup>#</sup>** |  |  |  |  |  |  |  |
| sMRI | 0.29 | 0.87 | 0.17 | 0.93 | 2.2 | 0.82 | 0.578 |
| GMA | 0.07 | 0.86 | 0.13 | 0.92 | 1.7 | 0.98 | 0.514 |
| HINE | 0.43 | 0.81 | 0.17 | 0.84 | 2.3 | 0.70 | 0.620 |
| sMRI plus GMA | 0.04 | 0.98 | 0.13 | 0.92 | 1.5 | 0.99 | 0.506 |
| sMRI plus HINE | 0.11 | 0.95 | 0.18 | 0.92 | 2.3 | 0.94 | 0.531 |

\*Predicts success perfectly due to 100% specificity
\*\*Predicts success perfectly due to 100% sensitivity
<sup>#</sup> GMFCS Level I is defined as an infant's ability to move in and out of sitting and floor sit with both hands free to manipulate objects. Infants crawl on hands and knees, pull to stand and take steps holding on to furniture. Infants walk 10 steps independently, with hands free, but with some gait abnormalities – includes toe walking, asymmetric walking, wide based gait with coordination or ataxic gait. Level II to V are characterized by increasing levels of limitations to walking and voluntary control of movement in other muscles.<sup>36,37</sup>
Abbreviations: CP= cerebral palsy, sMRI=structural magnetic resonance imaging, GMA=general movement assessment, HINE= Hammersmith Infant Neurological Examination, GMFCS= Gross Motor Functional Classification Scale

The individual post-test probabilities for varying levels of GMFCS in children with CP demonstrates considerable discrepancies in test accuracy for predicting level I from level II-V CP across various individual prognostic tests and their combinations (eTable 2). When test results are negative, the individual level probability of developing level I CP was like the cohort prevalence (i.e., pre-test probability) of level I CP (8.3%). Conversely, abnormal GMA results individually or when combined with abnormal sMRI, exhibited more than 80% probability for developing CP by age 2.

## Discussion

Our regional multisite prospective cohort study of infants born at ≤32 weeks’ GA assessed the predictive value of combining TEA-sMRI with GMA and/or HINE at 3-4 months CA for early CP prediction. The combined tests showed high specificity but suboptimal sensitivity in predicting CP, particularly for level I CP, representing most infants with CP in our cohort. Our findings offer new insights to guide the growing clinical practice of using sMRI with GMA/HINE for early prediction/diagnosis of CP.

While the 98% specificity of GMA in our cohort aligns with previous research, the 27% sensitivity is notably lower than the >95% sensitivity reported in most studies.^12,15–21,23,24,26,38–42^ These studies, including systematic reviews and meta-analyses,^12,43^ focused on high-risk infants with severe neuropathology, like perinatal asphyxia and stroke, and consequently a higher CP prevalence (6.3-52.4%), which may bias and inflate the test’s prognostic properties.^35^ Prechtl et al. reported high sensitivity by studying only infants with abnormal cranial ultrasound, resulting in a high CP prevalence of 37.6%.^39^ Cohort studies from unselected population-based samples, like ours, demonstrated much lower GMA sensitivity. A Swiss cohort study of very preterm infants (n=535) with a CP prevalence of 7% reported 56% sensitivity (95% CI: 38-72%).^26^ Similarly, a Dutch study from six well-baby clinics (n=455) reported 67% sensitivity (95% CI: 13–98%) in predicting CP.^44^

Other studies illustrating higher sensitivity in preterm infants (85-97%) often had smaller sample sizes (79-115)^40–42^ or earlier outcome assessments.^15,26,41,42^ An early diagnosis of CP before two years of age may overlook cases of CP with lower GMFCS levels and inflate the frequency of children with higher GMFCS levels, potentially leading to an increased sensitivity of GMA.^45^ Notably, our study also observed a higher sensitivity of GMA for predicting levels II-V CP (89%, Table 5).

Similarly, the 84% specificity of HINE in our study aligns with findings from previous studies; however, the 58% sensitivity was notably lower than prior investigations.^24,46–49^ Similar to GMA prognostic studies, most prior HINE studies^14,24,46–48,50^ examined high-risk infants exhibiting a higher prevalence of brain abnormalities and/or CP than generally observed in preterm infants.

While our study’s 90% specificity for brain MRI abnormalities aligned with previous findings, the 46% sensitivity was markedly lower than those reported by Novak et al. (86% to 89%).^16^ However, their data were derived from a cross-sectional MRI study,^51^ which cannot determine prognostic test properties, and from a systemic review that found only one eligible cohort study of 61 preterm infants.^17^ Prospective longitudinal studies in preterm infants,^10,52–54^ demonstrated between 50-67% sensitivity for moderate to severe abnormality on MRI at TEA to predict CP. The two largest MRI studies in preterm infants (total N=956)^25,53^ yielded a sensitivity of 48% and 60% for CP prediction, mirroring our findings and suggesting the need for additional tests to accurately screen for early CP.

Our study showed low sensitivity and high specificity for the combined use of sMRI, GMA, and/or HINE, suggesting it may not be adequate for screening very premature infants. While both high sensitivity and specificity are ideal for a prognostic test, high sensitivity is particularly crucial for effective screening.^55^ Due to the low sensitivity observed in our sample, even infants with negative results from combined assessments may still benefit from continued close monitoring for signs and symptoms of CP until at least 2 years CA. The low sensitivity of sMRI, alone or combined, may be partly due to the exclusion of some brain abnormalities, such as intraventricular hemorrhage without ventriculomegaly, that are predictive of CP in some studies but not consistently. However, this approach may improve sensitivity but at the cost of reducing specificity.

Our findings align with a multicenter cohort study of very premature infants (N=535), where combining cranial ultrasound abnormalities with GMA yielded only 15% (95% CI: 6-31%) sensitivity for detecting CP.^26^ Contrastingly, a retrospective case-control study reported 98% sensitivity and 99% specificity when combining neuroimaging, GMA, and HINE.^14^ Our results may differ due to study design. Case-control studies often involve artificially altered outcome prevalence (50% in 1:1 matching) rather than reflecting natural disease prevalence.^56^ Morgan et al., reported a 33% CP prevalence with 1:2 matching,^14^ considerably higher than the reported prevalence in very preterm infants, likely inflating sensitivity and specificity estimates.

Similarly, a small cohort study reported higher sensitivity (70%) for the sMRI and GMA combination,^15^ but their CP assessment was at 18 months CA. Because children with GMFCS I may be harder to diagnose before age 2, earlier diagnosis likely inflates the proportion of infants with higher GMFCS levels. In our study, the combined sensitivity of sMRI plus GMA to predict CP GMFCS levels II-V was 78%, with specificity of 99.7%, underscoring the robustness of this combined approach in identifying higher GMFCS levels. Interestingly, 72% of children with CP in our sample were classified as GMFCS I (n=28), who are more likely to score within typical ranges on the HINE^47^ and GMA,^57^ leading to false-negative results when assessed with the standard battery of early CP detection tools. This high proportion of children with GMFCS I in our sample may explain the lower sensitivity of these traditional early detection methods. Our findings emphasize the need to enhance detection methods for children at GMFCS level I, as infants in this group (with higher functional levels) respond particularly well to early therapeutic interventions.^58^

We demonstrated consistently high specificity for CP prediction, particularly when combining the assessment tools. When sMRI was combined with GMA, the specificity and PPV both reached 100%. This enables clinicians to confidently diagnose CP as early as 3-4 months CA when both tests are positive, rather than simply categorizing an infant as high-risk for CP. Early diagnosis can facilitate enrollment in CP prevention/treatment trials, prompt targeted and high-dose early intervention, streamline appropriate funding, and provide social support to their caregivers.

Our study highlights a notable difference in the sensitivity of GMA and HINE whether used alone (27% vs. 58%) and or incorporated with sMRI (22% vs. 32%). The additional difference in specificity suggests these tests provide distinct prognostic information and should not be used interchangeably. Our earlier research bolsters these findings, demonstrating that GMA and HINE independently relate to sMRI abnormalities and have limited correlation.^27^ This may be attributed to the likelihood that GMA and HINE assess distinct brain regions, indicating the importance of their combined utilization in clinical assessment.^27^

Our study’s strengths include being the largest prospective cohort study of preterm infants investigating the collective ability of sMRI, GMA, and HINE in forecasting CP. We minimized selection bias through consecutive recruitment of all preterm births born at ≤32 weeks’ GA in Greater Cincinnati. We performed sMRI at TEA and GMA between 12-18-weeks CA, the optimal time for test performance. HINE and GMA were conducted on the same day by one provider with high reliability, masked to clinical history and sMRI results. Similar rigor was applied to sMRI readings. However, some assessments involved multiple raters to confirm findings, and about one-third of outcome assessors at age 2 were not blinded to clinical history or prior test results, potentially inflating prognostic test properties, though this was not observed for test sensitivity. Additionally, 14% attrition may introduce bias, though baseline characteristics were similar between those who returned for follow-up and those who did not, except for GA. Lastly, we did not derive motor optimality scores from GMA videos, which may yield more accurate results than assessing only absent, sporadic, and abnormal fidgety movements.^59^

## Conclusion

Our prospective multisite cohort study found that combining sMRI and GMA exhibited high specificity for predicting CP, enabling confident early diagnosis of CP for abnormal test results. However, test sensitivity, particularly for children with CP GMFCS level I, remained suboptimal. Consequently, continued follow-up in high-risk clinics and/or early intervention programs is required for all very preterm infants until more sensitive tools, including quantitative MRI and machine learning prognostic models, are developed to improve early diagnosis and aggressive prevention/treatment of CP.

## Data Availability

All data produced in the present study are available upon reasonable request to the corresponding author.

## Acknowledgements

This research was supported by grants R01-NS094200 and R01-NS096037 from the National Institute of Neurological Disorders and Stroke (NINDS) and R01-HL164420 from the National Heart, Lung, and Blood Institute (NHLBI) of the National Institutes of Health. We sincerely thank the Cincinnati Infant Neurodevelopment Early Prediction Study (CINEPS) Investigators; Principal Investigator: Nehal A. Parikh, DO, MS; Collaborators (in alphabetical order): Mekibib Altaye, PhD, Anita Arnsperger, RRT, Traci Beiersdorfer, RN BSN, Kaley Bridgewater, RT(MR) CNMT, Tanya Cahill, MD, Kim Cecil, PhD, Kent Dietrich, RT, Christen Distler, BSN RNC-NIC, Juanita Dudley, RN BSN, Brianne Georg, BS, Cathy Grisby, RN BSN CCRC, Lacey Haas, RT(MR) CNMT, Karen Harpster, PhD, OT/RL, Lili He, PhD, Scott K. Holland, PhD, V.S. Priyanka Illapani, MS, Kristin Kirker, CRC, Julia E. Kline, PhD, Beth M. Kline-Fath, MD, Hailong Li, PhD, Matt Lanier, RT(MR) RT(R), Stephanie L. Merhar, MD MS, Greg Muthig, BS, Brenda B. Poindexter, MD MS, David Russell, JD, Sara Stacey, BSN, Kari Tepe, BSN RNC-NIC, Leanne Tamm, PhD, Julia Thompson, RN BSN, Jean A. Tkach, PhD, Hui Wang, PhD, Hui Wang, PhD, Jinghua Wang, PhD, Brynne Williams, RT(MR) CNMT, Kelsey Wineland, RT(MR) CNMT, Sandra Wuertz, RN BSN CCRP

## Data Sharing Statement

All deidentified participant data can be requested from the corresponding author upon reasonable request.

## Paper Presentation Information

This paper was presented at the Pediatric Academic Societies international meeting on April 30, 2023, in Washington, D.C. USA.

