## Supplement 1 for "Early Diagnosis of Cerebral Palsy in Preterm Infants with MRI, General Movements and Neurological Exam"

**eMethods**

**eTables**

**eReferences**

This supplementary material has been provided by the authors to give readers additional information about their work.

**eMethods**

*Brain MRI acquisition*

Infants being cared for at Cincinnati Children’s Medical Center, the sole imaging site, were imaged as inpatients and those born at the other four NICUs were imaged as outpatients after they were discharged from the NICU. Infants on positive pressure respiratory support were accompanied by a NICU nurse, respiratory therapist, and a neonatologist. All infants were scanned without sedation, using a feed and bundle approach, and fitted with ear plugs and muffs to protect hearing. We performed the following sequences on all infants: 3D T1-weighted Magnetization Prepared Rapid Acquisition Gradient Echo (MPRAGE): echo time (TE) = 3.4; repetition time (TR) = 7.3; inversion time (TI) =1110 ms; flip angle (FA) 11°, resolution 1.0 x 1.0 x 1.0 mm; 2:47min. T2-weighted MRI: TE/TR 166/10000 ms; FA 90°, resolution 1.0 x 1.0 x 1.0 mm; 3:53min. Sagittal susceptibility-weighted imaging: TE/TR 5.9/29 ms; FA 17°, resolution 0.6 x 0.6 x 1.5 mm; 3:27min.

*Neurodevelopmental outcome assessment:*

General Movement Assessment (GMA): GMA is an observational measure which categorizes the spontaneous infant movements in a quiet, alert infant. We video recorded infant general movements for 5 minutes between 12-18 weeks corrected age (CA) in the clinic using Prechtl methodology. ^1,2^ A single Prechtl advanced GMA certified examiner (KH), masked to clinical history and sMRI findings, scored the GMAs within 72 hours.^2^ The same examiner assessed the intra-rater reliability by re-evaluating 30 randomly selected videos 6 months later. Infants with normal fidgety movements exhibited variable, complex, and multiplanar movements, while those with absent or sporadic (≤3 seconds) fidgety movements were classified as having abnormal GMA. An absence of fidgety movements seen between the 2^nd^ and 4^th^ month of life is predictive of motor dysfunction especially cerebral palsy (CP).^2^ No cohort infants exhibited abnormal fidgety movements (exaggerated amplitude or speed).^3,4^ A General Movements Trust trainer (CP), blinded to clinical history, provided secondary assessment for equivocal videos.

Hammersmith Infant Neurological Examination (HINE): The HINE is a standardized neurologic examination for infants aged 2-24 months CA comprising of 26 items across 5 domains evaluating infants’ posture, tone, cranial nerves, reflexes and spontaneous and voluntary movements. ^5^ Each item is scored on a scale of 0-3, with domain scores summed for an overall HINE score ranging from 0 to 78. Scores <56 were considered abnormal.^6^ A single examiner (KH), masked to clinical history and MRI findings, performed HINEs on the same day as GMAs. The same examiner assessed the intra-rater reliability by retesting 15 randomly selected infants 2-4 weeks after their initial HINE.

Neurological examination to determine CP diagnosis: Certified examiners^7^ performed the Amiel-Tison standardized neurological exam^8^ at 22 to 26 months CA to assess motor function and diagnose CP using the Neonatal Research Network criteria: (1) abnormalities in tone, deep tendon reflexes, coordination, and/or movement, except isolated hypotonia or toe walking without tight ankles; (2) delays in motor milestones with a motor function disorder and (3) possible aberrations in reflexes and postural reactions. Because this definition permits a diagnosis of level I CP for a child with isolated toe walking with tight ankles, we required at least one additional delay in milestones or tone abnormalities for diagnosis of CP. We used the Gross Motor Function Classification System (GMFCS)^9,10^ to rate CP gross motor functional ability on a scale of I to V. Level I at 22 to 26 months CA is defined as an infant’s ability to move in and out of sitting and floor sit with both hands free to manipulate objects. Infants crawl on hands and knees, pull to stand and take steps holding on to furniture. Infants walk 10 steps independently, with hands free, but with some gait abnormalities – includes toe walking, asymmetric walking, wide based gait with coordination or ataxic gait. Level II to V are characterized by increasing levels of limitations to walking and voluntary control of movement in other muscles. Our primary outcome was diagnosis of CP at 22 to 26 months CA.

*Statistical analysis*

We analyzed baseline group differences in infants with and without CP and children who returned for follow-up at 2 years CA vs. those that who did not using Student’s t-test, Mann Whitney U, Chi-Squared test, or Fisher’s exact test as appropriate. Intra-rater reliability was assessed using kappa statistics for dichotomous variables and intraclass correlation coefficient for continuous variables. We used Fisher’s exact test to examine sensitivity, specificity, positive predictive value (PPV), and negative predictive value (NPV) in order to evaluate the predictive capabilities of the three tests individually and in combination for CP diagnosis at 22-26 months CA. Because PPV and NPV are affected by disease prevalence and not generalizable between centers, we calculated the likelihood ratio for a positive test (LR+) by dividing the sensitivity by 1-specificity and LR for a negative test (LR-) by dividing 1-sensitivity by the specificity. We used logistic regression to calculate if the odds ratio for the tests individually and in combination would be different for children with and without CP. Predicted post-test probabilities are usually easier to understand than the coefficients or the odds ratios. Thus, we employed the *margins* command to obtain predicted probabilities of different GMFCS levels in children with CP for each test and their combinations. We used Stata 17.0 (Stata Corp, College Station, TX, USA) for analysis. Two-sided P values <.05 were considered statistically significant.

**eTables**

**eTable 1.** Cerebral palsy diagnosis at 2 years corrected age (CA) and associated Gross Motor Functional Classification Scale (GMFCS) for all 8 infants diagnosed with abnormal structural MRI (sMRI) AND abnormal general movements assessment (GMA) and all 17 infants diagnosed with abnormal sMRI and abnormal Hammersmith Infant Neurological Exam (HINE) in the cohort.

|  | **sMRI Abnormal** | **GMA – Abnormal** | **HINE Abnormal** | **HINE Score** | **CP at 2 Years CA** | **GMFCS Level** |
| --- | --- | --- | --- | --- | --- | --- |
| **sMRI *and* GMA Abnormal** |  |  |  |  |  |  |
| Yes | Yes | Yes | Yes | 41 | Yes | IV |
| Yes | Yes | Yes | Yes | 51 | Yes | III |
| Yes | Yes | Yes | Yes | 30.5 | Yes | V |
| Yes | Yes | Yes | Yes | 52 | Yes | III |
| Yes | Yes | Yes | Yes | 37 | Yes | IV |
| Yes | Yes | Yes | Yes | 32 | Yes | V |
| Yes | Yes | Yes | Yes | 42.5 | Yes | V |
| Yes | Yes | Yes | Yes | 53.5 | Yes | I |
| 8/8 (100%) | 8/8 (100%) | 8/8 (100%) | 8/8 (100%) | 42.5  (30.5-53.5)* | 8/8 (100%) | N/A |
| **sMRI *and* HINE Abnormal**^¶^ |  |  |  |  |  |  |
| Yes | Yes | Yes | Yes | 42 | Yes | IV |
| Yes | Yes | Yes | Yes | 51 | Yes | III |
| Yes | Yes | No | Yes | 55 | No | N/A |
| Yes | Yes | No | Yes | 51.5 | Yes | I |
| Yes | Yes | Yes | Yes | 30.5 | Yes | V |
| Yes | Yes | No | Yes | 40.5 | No | N/A |
| Yes | Yes | No | Yes | 36 | Yes | III |
| Yes | Yes | No | Yes | 39.5 | No | N/A |
| Yes | Yes | Yes | Yes | 52 | Yes | III |
| Yes | Yes | Yes | Yes | 37 | Yes | IV |
| Yes | Yes | No | Yes | 49.5 | Yes | I |
| Yes | Yes | Yes | Yes | 32 | Yes | V |
| Yes | Yes | Yes | Yes | 42.5 | Yes | V |
| Yes | Yes | No | Yes | 51 | No | N/A |
| Yes | Yes | No | Yes | 46.5 | No | N/A |
| Yes | Yes | Yes | Yes | 53.5 | Yes | I |
| Yes | Yes | Missing | Yes | 45 | Yes | II |
| 17/17 (100%) | 17/17 (100%) | 8/16 (50.0%) | 17/17 (100%) | 45  (30.5-55)* | 12/17 (70.6%) | N/A |

^¶^Results for sMRI and GMA OR sMRI and HINE were identical to this group

*Median (IQR)

**eTable 2.** Individual predicted probabilities (in percentage and their 95% confidence intervals) for different severity levels of cerebral palsy (CP) based on three prognostic tests and their combinations.

| **Test** | **No CP** | **Level I CP^#^** | **Levell II-V CP^#^** |
| --- | --- | --- | --- |
| ***Negative test*** |  |  |  |
| sMRI | 92.9 (89.9, 95.8) | 5.4 (3.0, 7.8) | 1.7 (0.5, 2.9) |
| GMA | 91.5 (88.4, 94.6) | 7.2 (4.5, 10.0) | 1.3 (0.2, 2.4) |
| HINE | 94.0 (91.1, 96.8) | 4.7 (2.3, 7.0) | 1.4 (0.3, 2.4) |
| sMRI and GMA | 91.0 (87.8, 94.1) | 8.4 (5.4, 11.5) | 0.6 (-0.2, 1.4) |
| sMRI and HINE* | 91.8 (88.8, 94.8) | 7.0 (4.3, 9.7) | 1.2 (0.2, 2.3) |
| ***Positive test*** |  |  |  |
| sMRI | 59.1 (44.7, 73.4) | 27.4 (16.8, 38.0) | 13.6 (4.9, 22.2) |
| GMA | 16.4 (-0.1, 32.9) | 42.2 (24.9, 59.4) | 41.5 (15.1, 67.9) |
| HINE | 66.4 (55.0, 77.7) | 23.8 (14.7, 33.0) | 9.8 (3.5, 16.1) |
| sMRI and GMA | 0.9 (-1.2, 3.0) | 11.5 (-9.4, 32.5) | 87.6 (64.9, 110.3) |
| sMRI and HINE* | 15.2 (0.5, 29.9) | 41.4 (24.4, 58.3) | 43.5 (18.8, 68.1) |

*Results for sMRI and GMA OR sMRI and HINE were identical to this group

^#^ GMFCS Level I is defined as an infant’s ability to move in and out of sitting and floor sit with both hands free to manipulate objects. Infants crawl on hands and knees, pull to stand and take steps holding on to furniture. Infants walk 10 steps independently, with hands free, but with some gait abnormalities – includes toe walking, asymmetric walking, wide based gait with coordination or ataxic gait. Level II to V are characterized by increasing levels of limitations to walking and voluntary control of movement in other muscles.^9,10^

Abbreviations: CP= cerebral palsy, sMRI=structural magnetic resonance imaging, GMA=general movement assessment, HINE= Hammersmith Infant Neurological Examination, GMFCS= Gross Motor Functional Classification Scale
